# Exosomal miRNAs from maternal vaginal discharge as biomarkers for preterm labor: non-invasive liquid biopsy

**DOI:** 10.1101/2024.04.03.24304152

**Authors:** Taewoon Kim, Jee Yoon Park, Hyo Jin Lee, Bo young Choi, Hyeon Ji Kim, Luke P. Lee, Jong Wook Hong

**Affiliations:** Department of Bionanotechnology, Graduate School, Hanyang University, Seoul 04763, Korea; Department of Obstetrics and Gynecology, Seoul National University Bundang Hospital, Seongnam, Korea; Department of Obstetrics and Gynecology, Seoul National University College of Medicine, Seoul, Korea; Harvard Medical School, Harvard University; Department of Medicine, Brigham and Women’s Hospital, Boston, Massachusetts, USA; Department of Bioengineering, University of California at Berkeley, Berkeley, CA, USA; Department of Electrical Engineering and Computer Science, University of California at Berkeley, Berkeley, CA, USA; Department of Biophysics, Institute of Quantum Biophysics, Sungkyunkwan University, Suwon, Korea; Department of Chemistry & Nanoscience, Ewha Womans University, Seoul, Korea; Department of Medical and Digital Engineering, Graduate School, Hanyang University, Seoul 04763, Korea; Department of Bionanoengineering, Hanyang University, Gyeonggi-do 15588, Korea

**Keywords:** preterm labor, vaginal discharge, extracellular vesicles, exosomes, RNA-seq

## Abstract

Preterm labor is a serious issue that can lead to preterm birth, posing significant risks to both the mother and the neonate. Despite the high incidence of approximately 15 million preterm births worldwide per year, there is a lack of sufficient strategies for predicting and preventing preterm labor. Here, we found that exosomal miRNAs in maternal vaginal discharge can serve as biomarkers for early diagnosis of life-threatening conditions in both the mother and neonate. Our non-invasive biopsy of vaginal discharge using a swab allows us to isolate enriched exosomes via an advanced microfluidic platform called BEST (Biologically intact Exosome Separation Technology). We have identified specific miRNAs differentially expressed in mothers with preterm labor compared to those with full-term mothers. These miRNAs included hsa-miR-206, which was up-regulated in preterm labor, and hsa-miR-3674, hsa-miR-365a-5p, and hsa-miR-193b-3p, which were down-regulated. We believe our discovery of unique miRNAs as biomarkers can aid in early detection and effective treatment of preterm labor, potentially revolutionizing global healthcare.

## Main

Preterm birth, defined as delivery before 37 weeks of gestation^1,2^, poses a profound global health challenge, with prevalence rates ranging from 5% in Europe to 18% in Africa^3^. The estimated worldwide occurrence of preterm birth was approximately 10% in 2020, solidifying its status as the leading cause of neonatal morbidity and mortality^4^. Despite an annual occurrence of approximately 15 million preterm births worldwide, practical strategies for prediction and prevention are still insufficient^5,6^. Infants born prematurely not only encounter heightened risks during birth and infancy, but also contend with enduring health issues throughout their lives, including conditions such as cerebral palsy, cognitive disability, epilepsy, blindness, or hearing loss^3,7^. When stratifying by degrees of prematurity, the risks are highest for babies born extremely preterm before 28 weeks gestation, followed by those born very preterm between 28 and less than 32 weeks, and moderate to late preterm between 32 and less than 37 weeks^8-10^. This underscores the pressing need for further research and comprehensive public health initiatives.

The causes of spontaneous preterm birth are preterm labor, preterm premature rupture of membranes, and acute cervical insufficiency^11,12^. Among those, preterm labor is defined as the development of regular uterine contractions before the gestational period reaches 37 weeks, and cervical remodeling, such as dilatation or progression of effacement, usually accompanies resulting in preterm birth^13^. Two-thirds of all preterm births have been known to result from spontaneous preterm labor, and most pregnant women with preterm labor suffer various degrees of abdominal pain and/or back pain due to increased uterine contractility^14^. Moreover, uncontrolled preterm labor ultimately results in gross premature rupture of membranes. Tocolytic agents to relieve uterine contractility are used to manage preterm labor; however, global guidelines or meta-analyses have suggested they are not significantly effective in reducing the rate of preterm birth^15-17^. Therefore, tocolytics are recommended to be administered only for a short time for antenatal corticosteroids to accelerate fetal lung maturation or for transferring the patient to tertiary centers with neonatal intensive care units^18^. There are several pieces of evidence that intraamniotic microbial invasion with inflammation caused by ascending infection is the most critical pathophysiology of spontaneous preterm birth; antibiotics are not recommended to be used for preterm labor with intact membranes^19,20^.

Exosomes have become a focal point in biomedical research due to their crucial influence on various biological functions and their ability to modulate microenvironments^21,22^. Exosomes, classified as membrane-bound nanovesicles with sizes ranging from 30 to 200 nm^23^, are released from diverse cell types and bodily fluids, including blood^24^, saliva^25^, breast milk^26^, urine^27^, tears^28^, and vaginal discharge^29^. These small vesicles transport bioactive compounds like proteins, nucleic acids, and lipids^30,31^. In the specific context of pregnancy, current research highlights the significant impact of exosomes as potential mediators in maternal-fetal communication^32,33^. During pregnancy, intricate communication between maternal and fetal cells is vital for properly developing and maintaining the gestational environment. Moreover, exosomes derived from plasma have demonstrated promising diagnostic potential for various pregnancy outcomes, establishing a solid foundation for exploring their role in predicting complications related to preterm birth^34-36^.

Vaginal discharge is a composite of mucus originating from the vagina. It is consistently generated by secretory cells in the endocervix and the anterior vaginal epithelium. This mucus protects the female reproductive tract, ensuring the moisture and lubrication of epithelial surfaces to safeguard against potential threats^37-39^. Despite regular exposure to pathogenic microorganisms, the female genital tract maintains a relatively low incidence of infection, underscoring the presence of numerous defense mechanisms^40,41^. Critical components of this intricate defense system involve efficiently removing adherent bacteria through shedding epithelial cells^42^. Additionally, the crucial hydration of the cervical-vaginal mucosa is achieved through secretions from cervical and vaginal glands alongside plasma transudate^43^. These collective mechanisms work synergistically, protecting against potential threats and reinforcing the tract’s resilience in the face of microbial challenges. In recent studies, a paper has been published indicating the significant impact of exosomes derived from female vaginal discharge on the function and fertility of human sperm^44^.

In this study, we aim to elucidate the pathology of preterm labor through RNA-seq analysis of exosomes derived from vaginal discharge (VD) between patients with preterm labor (PTL) and asymptomatic term birth (TB). By scrutinizing exosome cargo from virginal discharge, we seek to unravel potential differences contributing to the onset of preterm labor. Ultimately, we aim to identify key molecular signatures that could serve as early diagnostic biomarkers for preterm labor and lay the groundwork for future clinical interventions in high-risk preterm birth scenarios.

Our workflow, illustrated in Fig. 1A–C, involved the initial dilution and centrifugation of VD samples to reduce viscosity and eliminate cell debris (Fig. S1). Subsequently, exosomes were isolated from pretreated VD samples using a biologically intact exosome separation technology (BEST) chip, which is the advanced version of our tunable microfluidics^45^ (Fig. 1B). Our BEST chip has been designed to separate exosomes while preserving their biological integrity. As a result, we can obtain the highest quality exosomes that provide the best results of exosomal miRNAs as biomarkers. Isolated exosomes were then used for downstream analysis, as shown in Fig. 1C. The study of particle size distribution in VD and exosome samples from PTL and TB revealed a significant finding: size peak shifting occurred from 155/225 nm to 65/145 nm following exosome separation, as shown in Fig. 1D and E. This finding underscores the effectiveness of our exosome isolation method. Moreover, a substantial increase in exosome concentration and protein content was noted in PTL compared with TB (Fig. S2A and B).

**Fig. 1.**
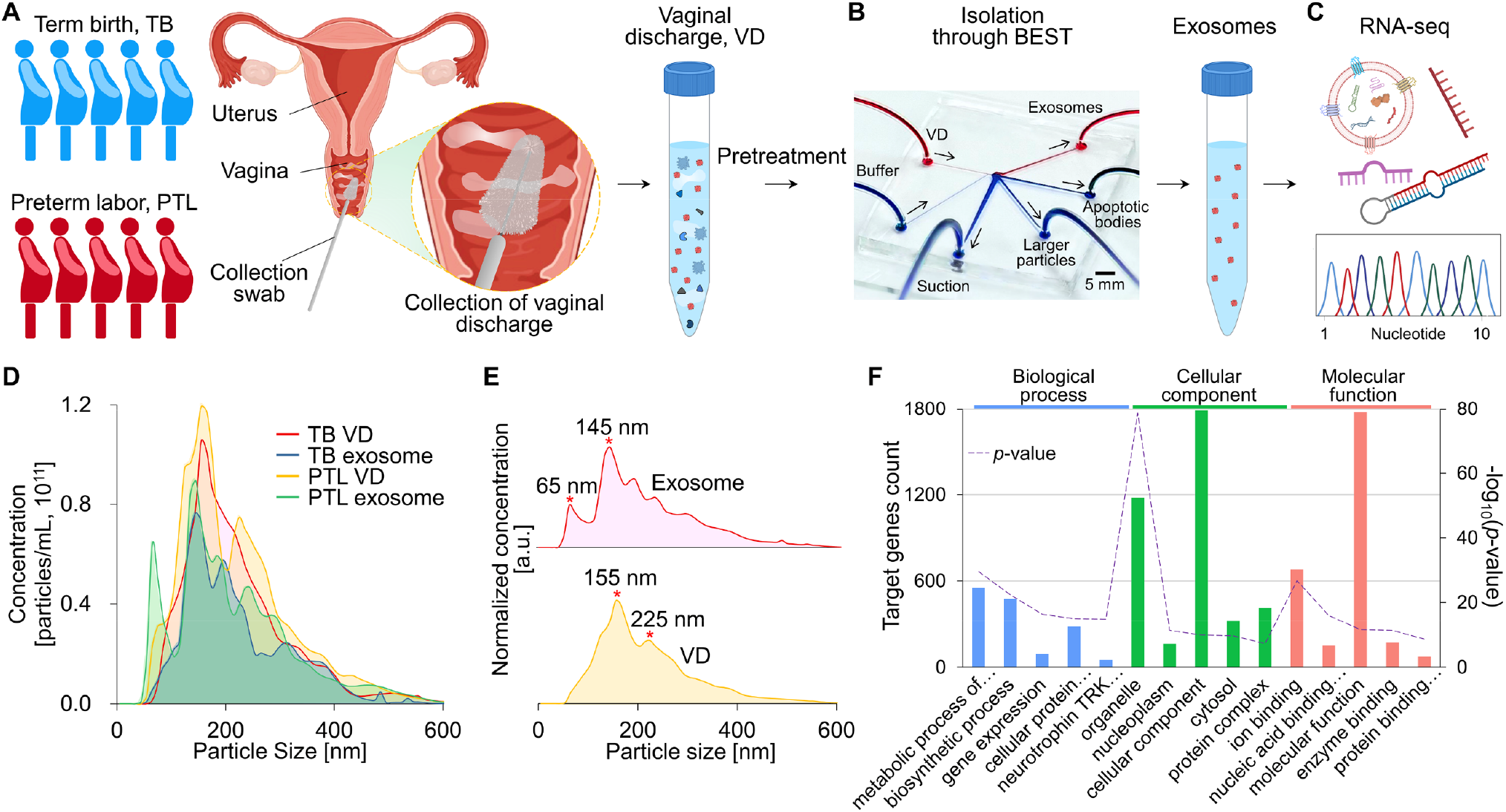
Overview of the analysis of exosomes from the vaginal discharge of pregnant women to diagnosis of preterm labor. (A) Collection of vaginal discharge (VD) using a swab from pregnant women with asymptomatic term birth (TB) and patients with preterm labor (PTL). (B) Isolation of exosomes using our biologically intact exosome separation technology (BEST). (C) Analysis of exosomal miRNA at the genomic level to identify biomarkers that can diagnose preterm labor. (D) Particle concentration and size distribution of VD and VD-derived exosomes from TB and PTL were measured through nanoparticle tracking analysis. (E) Comparison of particle size distributions before and after isolation of exosomes from VD. The peaks in size were marked after normalization. (F) Gene Ontology (GO) enrichment analysis of differentially expressed miRNA from VD-derived exosomes of TB and PTL. The top 5 most significant GO terms of miRNA target genes were listed in three categories: biological process, cellular component, and molecular function, respectively.

The analysis of exosomal miRNAs provided insights into the molecular signatures associated with PTL and TB. We performed Gene Ontology (GO) enrichment analysis and Kyoto Encyclopedia of Genes and Genomes (KEGG) pathway enrichment on the differentially expressed miRNAs between TB exosome and PTL exosome, following the criteria of Foldchange > 2.0, log_2_(Normalized data) > 1.0, and *p*-value < 0.05, to explore the potential biological functions of miRNAs (Fig. 1F, Fig. S3, and S4). The GO analysis revealed the top 5 most significant GO terms for biological process (BP), cellular component (CC), and molecular function (MF), respectively. The results indicated that BP primarily involves the metabolic process of cellular nitrogen compound, biosynthetic process, gene expression, cellular protein modification process, and neurotrophin tyrosine kinase (TRK) receptor signaling pathway. Regarding CC enrichment analysis, miRNAs significantly participated in the organelle, nucleoplasm, cellular component, cytosol, and protein complex. Regarding MF analysis, miRNAs were mainly enriched in ion binding, nucleic acid binding transcription factor activity, and molecular function, followed by enzyme binding and protein binding transcription factor activity. The KEGG pathway analysis of target genes revealed 17 significantly enriched pathways (*p*-value < 0.05), including biosynthesis of glycosphingolipid, proteoglycans in cancer, and glioma.

Furthermore, Venn diagram analysis revealed distinct miRNA profiles between TB VD, TB exosome, PTL VD, and PTL exosome groups. Overall, 1,437 different miRNAs were identified with 835, 849, 864, and 802 miRNAs from TB VD, TB exosome, PTL VD, and PTL exosome group, respectively, meeting the criterion of log_2_(Normalized data) ≠ 0. There were 414 common genes present in all four groups (Fig. 2A). The principal component analysis (PCA) plot indicated differentiation between PTL and TB samples. According to the three-dimensional (3D) plot, the samples were categorized into four groups (Fig. 2B). Each group appeared quite distinct, with the first three PCs explaining 39% of data variance (PC1 = 19.3%, PC2 = 12.9%, PC3 = 6.8%).

**Fig. 2.**
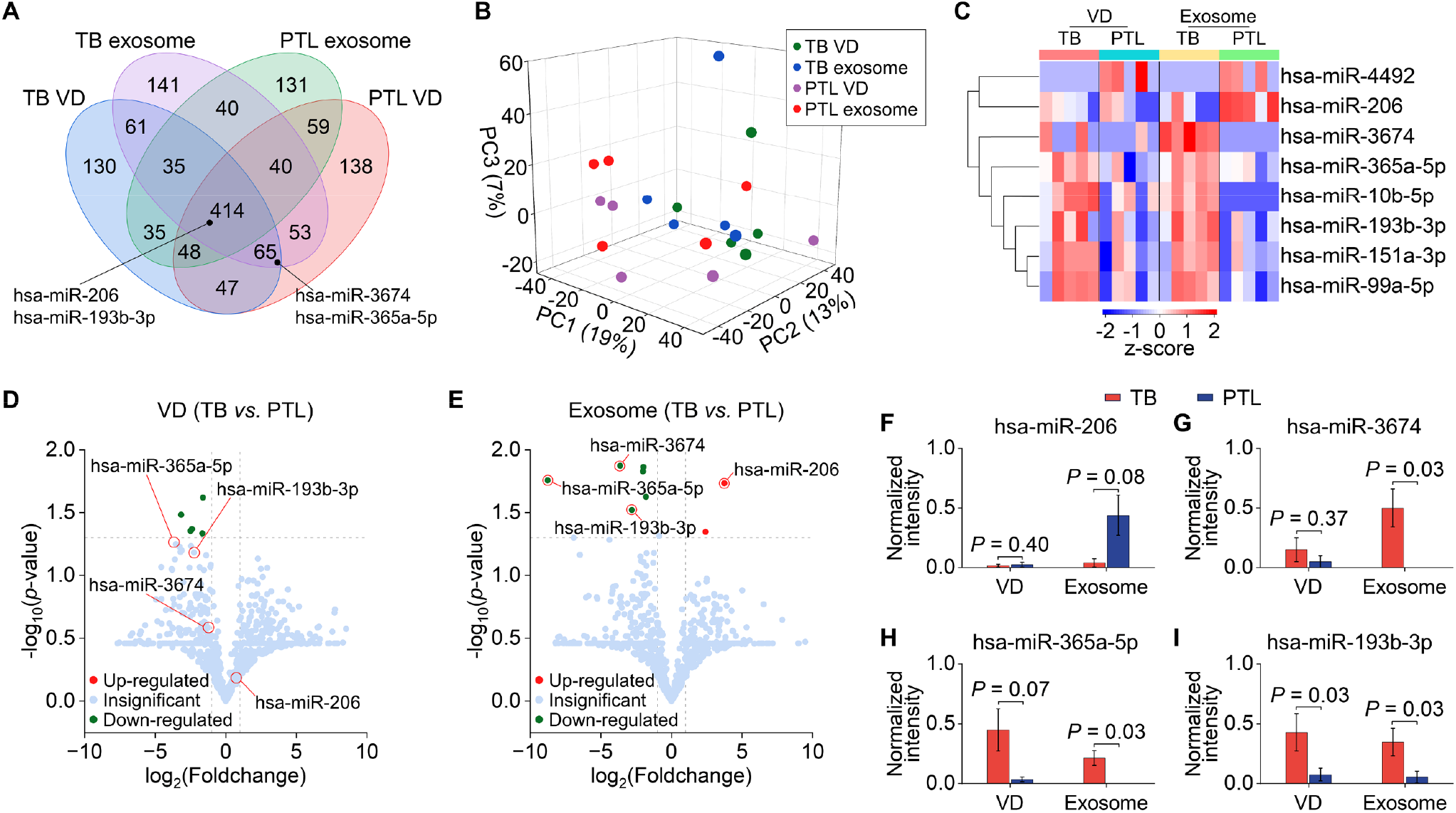
Identification of exosomal miRNA in maternal vaginal discharge as potential biomarkers for preterm labor. (A) Venn diagram of total genes from TB and PTL of VD and exosomes. TB, term birth; PTL, preterm labor; VD, vaginal discharge. (B) The PCA plot shows close clustering in each group. (C) Heatmap of the relative abundance of identified exosomal miRNA in the vaginal discharge and exosome of PTL and TB. (D) Volcano plot showing the distributions of differential expression miRNA between vaginal discharge of PTL and TB, and (E) exosome of PTL and TB. (F)–(I) The gene expression levels of the most differentially expressed four genes.

We further investigated the expression profiles of 8 miRNAs, which were differentially expressed in VD-derived exosomes, using criteria of Foldchange > 2.0, log_2_(Normalized data) > 1.0, and *p*-value < 0.05 (Fig. 2C). The hierarchical clustering heatmap in exosome groups indicated differentiation between PTL and TB compared to the VD group. To gain a deeper understanding of the impact of VD-derived exosomes in distinguishing between PTL and TB, we performed a volcano plot analysis to examine miRNA expression differences. Applying criteria of Foldchange > 2.0 and *p*-value < 0.05, a comparison of VD between PTL and TB revealed 6 down-regulated miRNAs, as shown in Fig. 2D. However, we identified 8 differentially expressed miRNAs in exosomes between PTL and TB. Among these, 2 miRNAs were up-regulated, and 6 miRNAs were down-regulated (Fig. 2E). Notably, key differential genes in exosomes, such as hsa-miR-206, hsa-miR-3674, hsa-miR-365a-5p, and hsa-miR-193b-3p, showed significance exclusively in exosomes, suggesting their potential association with PTL. Analyzing pooled miRNA expression data, we identified the four most differentially expressed miRNAs, with their profiles displayed in Fig. 2F–I. Specifically, hsa-miR-206 exhibited up-regulation, while hsa-miR-3674, hsa-miR-365a-5p, and hsa-miR-193b-3p were down-regulated in PTL compared to TB.

In this study, we conducted a comprehensive analysis of VD samples collected non-invasively from individuals experiencing PTL and those with TB, aiming to identify molecular signatures associated with the risk of PTL. Our findings highlight the potential utility of VD-derived exosomes as non-invasive biomarkers for distinguishing between PTL and TB cases. Through meticulous analysis, we observed a notable shift in particle size distribution following exosome isolation and a significant increase in exosome and protein concentration in PTL samples compared to TB. These observations underscore the effectiveness of our isolation method and the potential of exosome-based biomarkers in PTL diagnosis.

Our miRNA profiling provided valuable insights into the molecular landscape associated with PTL. Distinct miRNA profiles were delineated between PTL and TB groups, with PCA corroborating differentiation between the two cohorts. GO enrichment analysis revealed the involvement of miRNAs in critical biological processes, cellular components, and molecular functions relevant to preterm birth pathogenesis. Furthermore, differential expression analysis uncovered a panel of miRNAs exhibiting significant dysregulation in PTL compared to TB, particularly within exosome samples. Notably, miRNAs such as hsa-miR-206, hsa-miR-3674, hsa-miR-365a-5p, and hsa-miR-193b-3p have emerged as potential candidates associated with PTL. We explored the reported functions of these miRNAs, and a few studies investigated miRNAs in pregnancy. For example,

Deng *et al*. discovered that hsa-miR-206 plays a role in myometrium contraction using uterine tissue^46^. Akehurst *et al*. also found that has-miR-206 significantly increased in maternal plasma samples of pregnant women with preeclampsia^47^. Unlike those studies, our study involves research conducted solely using vaginal discharge, which is characterized as non-invasive samples that can be obtained quickly and repeatably as well. Since pregnancy research must encounter maternal and fetal stability, exploring biomarkers related to pregnancy outcomes from non-invasive methods is precious. We also found hsa-miR-193b-3p as a miRNA involved in cell differentiation and proliferation^48^. The association of hsa-miR-365a-5p with the oxytocin signaling pathway was identified through a miRNA target study conducted using miRWalk 3.0^49^. Given oxytocin’s critical functions in uterine contractions during parturition and milk release during lactation, there is a potential linkage between hsa-miR-365a-5p and PTL. However, information regarding the roles of hsa-miR-3674 in PTL remains to be limited based on our investigation.

The dysregulation of the four miRNAs, which were overexpressed in exosomes but appeared insignificant in the vaginal discharge itself, can be attributed to several factors. Firstly, vaginal discharge comprises a heterogeneous mixture of various RNA species and other molecules, potentially diluting the relative abundance of miRNAs. Additionally, the expression of miRNAs in vaginal discharge may fluctuate over time, exhibiting subtle changes under specific conditions or time points. Moreover, factors influencing the stability and preservation of miRNAs within vaginal discharge could also contribute to these observations. Therefore, careful consideration of these factors is necessary, and further experiments and analyses may be required to fully understand why the miRNAs overexpressed in exosomes exhibited insignificance in the vaginal discharge itself.

Despite the promising findings, several limitations should be acknowledged. Firstly, the sample size in our study was relatively small, emphasizing the need for validation in larger cohorts to ensure the reliability and applicability of our findings. Moreover, the cross-sectional design of our study restricts our ability to determine causality between altered exosomal profiles and PTL. Longitudinal studies tracking changes in exosomal markers throughout pregnancy could offer valuable insights into the predictive value of these biomarkers for PTL.

In conclusion, we conducted a study to determine if analyzing exosomal miRNAs from a non-invasive biopsy of maternal vaginal discharge could be an effective method for identifying preterm birth at an early stage. Our process involved using a swab to collect vaginal discharge, which is non-invasive and localized. We then used our BEST chip to obtain enriched exosomes and identify the molecular signatures associated with preterm labor. The ultimate goal is to develop a personalized precision molecular diagnostic platform to assess and manage the risk of preterm birth and significantly improve maternal and neonatal outcomes. However, further investigation involving a large population and long-term follow-up is necessary to confirm these results and determine the clinical significance of exosomal biomarkers in prenatal care and clinical practice in obstetrics.

## Methods

### Non-invasive and local collection of vaginal discharge

Vaginal discharge samples were obtained from five patients with preterm labor and five healthy controls with term gestational period (mean gestational age at sampling ± standard deviation, 29.8 ± 4.09 *vs*. 38.4 ± 0.84, respectively), following approval by the Seoul National University Bundang Hospital Institutional Review Board (B-2312-871-301). Preterm labor was defined as regular uterine contractions with cervical change or shortening of cervical length through transvaginal ultrasound but without evidence of ruptured membranes. All patients with preterm labor were hospitalized and received tocolytic agents. Informed consent was obtained from all participants. Through speculum examination, vaginal discharge samples were collected using a specimen collection swab (NobleBio, Republic of Korea) by obstetricians. They were then transferred to a transport medium (NobleBio, Republic of Korea) suitable for downstream analysis. They were stored at -80°C for subsequent analysis.

### Isolation of exosomes

Vaginal discharge underwent a 10-fold dilution in Dulbecco’s Phosphate-Buffered Saline (DPBS, SolBio, Republic of Korea) to decrease its viscosity (Fig. S1). The diluted sample was centrifuged at 2,000 × g for 30 minutes to eliminate whole cells and cell debris^50,51^. The resulting supernatant was then used for isolating exosomes derived from vaginal discharge using biologically-intact exosome separation technology, BEST^45^.

### Nanoparticle tracking analysis (NTA)

NTA measurements were conducted with the NanoSight LM10 (Malvern Instruments Ltd., UK). Before each measurement, the sample chamber was thoroughly cleaned with ethanol, ensuring optimal conditions for analysis. Due to the requirement of the NTA system for samples to fall within a concentration range of 10^6^ to 10^9^ vesicles per mL, all samples were subjected to a 1000-fold dilution to meet this specified concentration range. The sample was injected into the chamber using a syringe until the liquid overflowed from the opposite side. A thermal sensor facilitated temperature monitoring during each measurement to account for potential variations. NTA 3.1 software was employed for accurate data recording and analysis, enabling precise measurements of vesicle concentration and size distribution. Each sample was measured three times, lasting 15 seconds, and the sample chamber was meticulously cleaned with clean deionized water between each experiment to prevent any potential cross-contamination.

### Protein measurement

Samples were mixed with RIPA lysis buffer containing a 1x protease inhibitor cocktail and incubated for 10 min at 4°C. Subsequently, the lysates were centrifuged at 14,000 g for 10 min at 4°C to remove unlysed debris. Protein supernatants were then collected and utilized for a bicinchoninic acid (BCA) protein assay to determine protein concentrations. The assays were conducted following the manufacturer’s protocols.

### RNA extraction

Following the manufacturer’s protocol, exosomal RNA was extracted using TRIzol LS reagent (Ambion, Life Technology, USA). The quality of the RNA was evaluated using the RNA 6000 Pico Chip on an Agilent 2100 bioanalyzer (Agilent Technologies, Netherlands), while quantification was conducted utilizing the NanoDrop 2000 Spectrophotometer system (Thermo Fisher Scientific, USA).

### Library preparation and sequencing

Library construction was conducted using the NEBNext Multiplex Small RNA Library Prep kit (New England BioLabs, Inc., USA), following the manufacturer’s guidelines for both control and test RNAs. The process involved ligating adaptors to total RNA samples and synthesizing cDNA with adaptor-specific primers and reverse transcriptase. PCR was utilized for library amplification, followed by cleanup using the QIAquick PCR Purification Kit (Qiagen, Inc., Germany) and polyacrylamide gel electrophoresis (PAGE) gel. The yield and size distribution of the small RNA libraries were evaluated using the Agilent 2100 Bioanalyzer instrument with the High-sensitivity DNA Assay (Agilent Technologies, Inc., USA). High-throughput sequencing was performed using the NextSeq550 system with single-end 75 sequencing (Illumina, USA).

### Data analysis

Sequence reads were aligned using the bowtie2 software to generate a bam file, with mature miRNA sequences as the mapping reference. Bedtools v2.25.0^52^ and Bioconductor^53^, implemented in the R statistical programming language^54^, retrieved read counts mapped to the mature miRNA sequences from the alignment file. These read counts were then employed to assess the expression levels of miRNAs. For comparison between samples, the CPM+TMM normalization method was used. Gene Ontology (GO) enrichment analysis and Kyoto Encyclopedia of Genes and Genomes (KEGG) pathway enrichment were performed by DIANA^55^. All numerical graphs were generated using Origin software, Mircosoft Excel, and SRplot^56^.

### Statistical analysis

Statistical analysis was performed using Microsoft Excel, wherein descriptive statistics, including means and standard errors (S.E.), were computed for the dataset. The comparison of variables between two groups—namely healthy controls with term birth (n=5) and patients with preterm labor (n=5)— using the independent two-tailed t-test with a significance level of α = 0.05. Additionally, within each group, a paired t-test was employed to assess differences in variables. This combined use of independent and paired t-tests facilitated a comprehensive examination of both inter-group and intra-group variations, ensuring a thorough analysis of the collected data.

## Data Availability

All data produced in the present work are contained in the manuscript

## Acknowledgments

This research was supported by the National Research Foundation of Korea (NRF) grant funded by the Korean government (MSIT) (2022R1A2C1093134) and the Creative-Pioneering Researchers Program through Seoul National University (800-20230492).

