## Supplementary Materials for "Exosomal miRNAs from maternal vaginal discharge as biomarkers for preterm labor: non-invasive liquid biopsy"

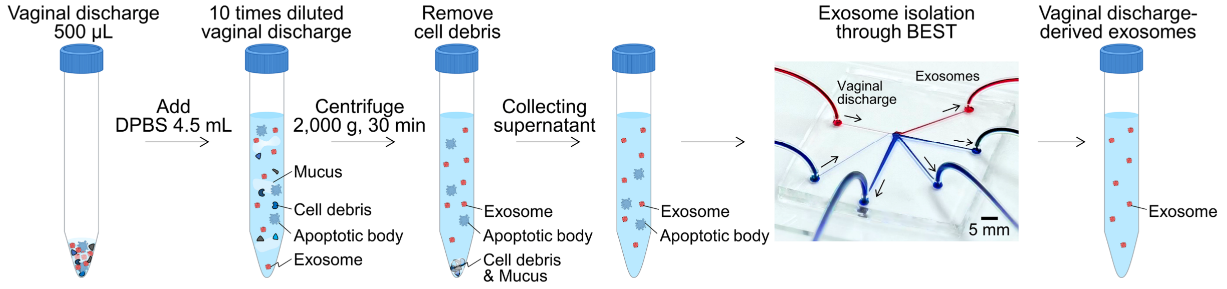


**Fig. S1 Vaginal discharge sample pretreatment process.** The viscosity of vaginal discharge was reduced by performing a 10-fold dilution in Dulbecco's Phosphate-Buffered Saline (DPBS). Subsequently, the diluted sample underwent centrifugation at 2,000 × g for 30 minutes to eliminate whole cells and cell debris, facilitating the isolation of vaginal discharge-derived exosomes using biologically intact exosome separation technology (BEST).

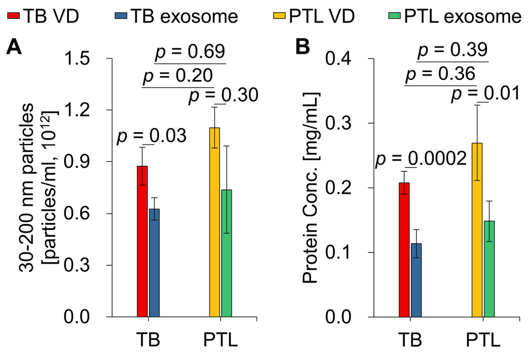


**Fig. S2 Characterization of exosomes derived from vaginal discharge (VD).** (A) Exosome-sized particle concentrations were compared between term birth (TB) and preterm labor (PTL), revealing higher concentrations in PTL. (B) Protein concentrations were compared between TB and PTL.

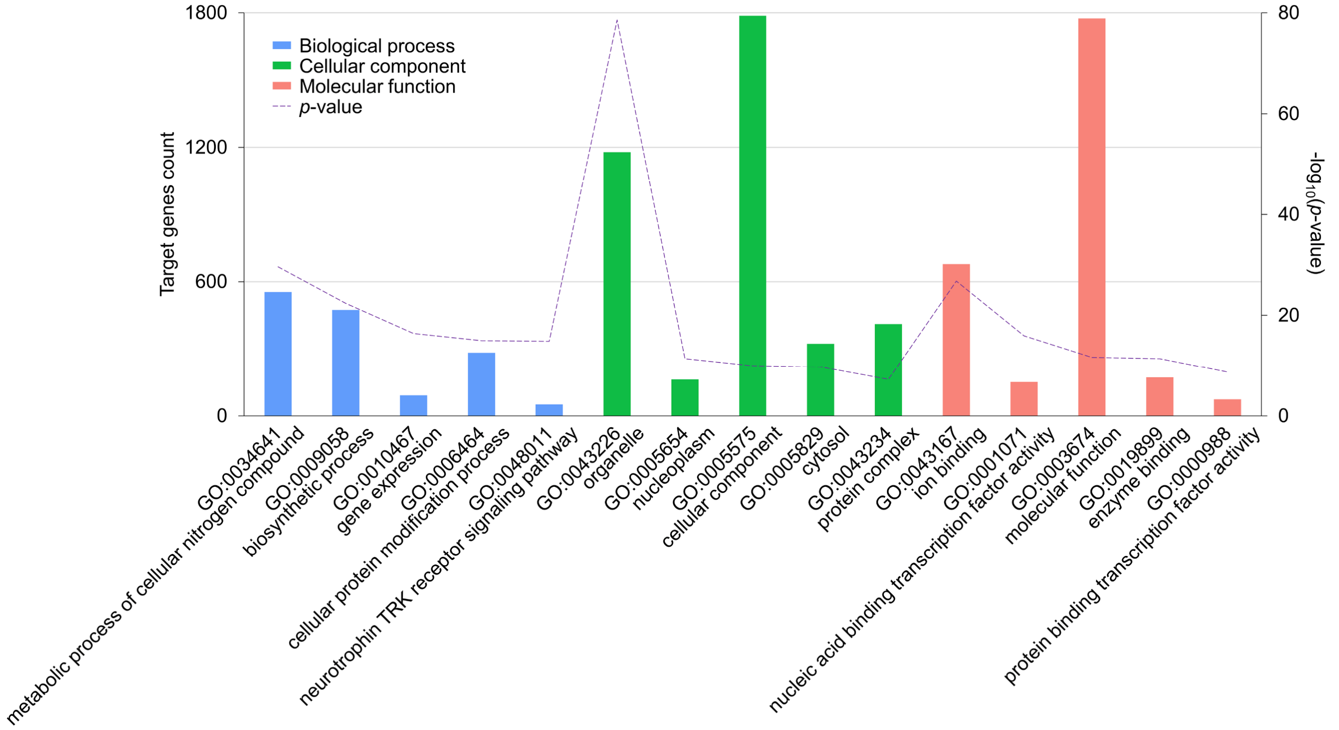


**Fig. S3 Gene Ontology (GO) enrichment analysis with uncropped annotations.**

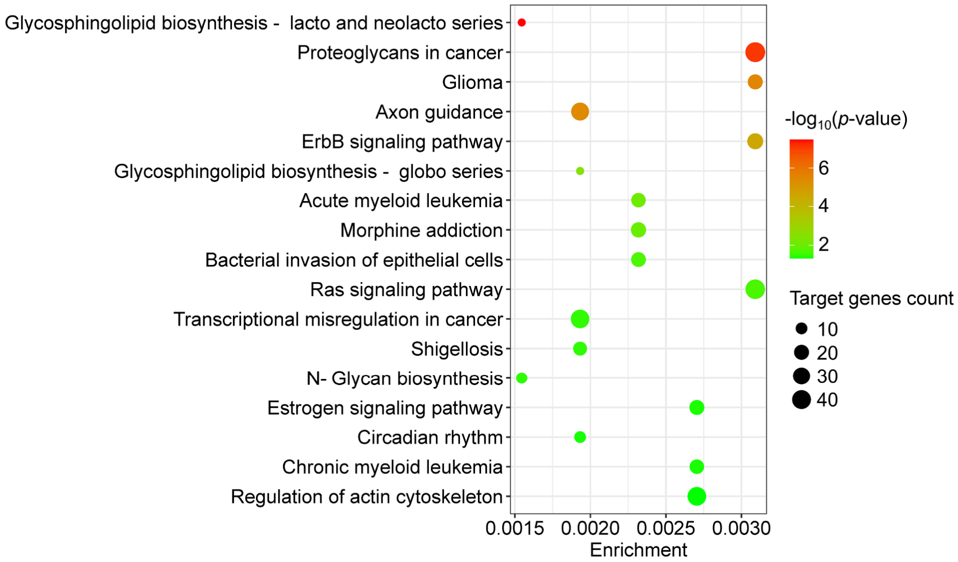


**Fig. S4 Scatter plot for KEGG pathway enrichment of miRNA target genes.** Enrichment is calculated as the ratio of differentially expressed miRNAs to all miRNAs annotated within a specific pathway term. Larger dots represent a higher count of target genes, and the red coloration indicates a more notable significance in terms of *p*-values. KEGG, Kyoto encyclopedia of genes and genomes.
